# Asymptomatic carriage of *Plasmodium falciparum* in children living in a hyperendemic area occurs independently of IgG responses but is associated with a balanced inflammatory cytokine ratio

**DOI:** 10.1101/2022.05.04.22274662

**Authors:** Balotin Fogang, Matthieu Schoenhals, Franklin M. Maloba, Marie Florence Biabi, Estelle Essangui, Christiane Donkeu, Glwadys Cheteug, Marie Kapen, Rodrigue Keumoe, Sylvie Kemleu, Sandrine Nsango, Douglas H. Cornwall, Carole Eboumbou, Ronald Perraut, Rosette Megnekou, Tracey J. Lamb, Lawrence S. Ayong

## Abstract

**Background:** Asymptomatic carriage of infected red blood cells (iRBCs) can be prevalent in communities regardless of transmission patterns and can occur with infection of different *Plasmodium* species. Clinical immunity dampens the inflammatory responses leading to disease symptoms in malaria. The aim of this study was to define the immunological correlates of asymptomatic carriage of *P. falciparum* in a highly exposed population.

**Methods:** 142 asymptomatic *Plasmodium*-infected individuals greater than 2 years of age without fever (body temperature <37.5°C) were followed weekly for 10 weeks before being treated with artemisinin-based combination treatment (ACT). Plasma levels of 38 cytokines were measured at baseline by Luminex and the quantity and growth inhibitory activities of circulating parasite-reactive antibodies measured. The *Plasmodium* antigen tested included *P. falciparum* merozoite extract (ME) and schizont extract (SE), and the recombinant proteins Erythrocyte binding antigen 175 (EBA-175) and merozoite surface protein 1 (MSP-1_19_).

**Results:** Median levels of IgG against *P. falciparum* EBA-175 and MSP-1_19_ at baseline were significantly higher in those older than 20 years of age compared with the younger age group and appeared to correlate with better parasite control. Amongst all participants there were no discernible changes in IgG levels over time. Parasite density was higher in the younger age group and associated with IL-10, TNF-α and MCP-1 levels. A balanced IL-10:TNF-α ratio was associated with asymptomatic malaria regardless of age, and balanced ratios of IL-10/TNF-α and IL-10/IFN-γ were the only significant correlate of maintenance of asymptomatic malaria over the course of the study in individuals 20 years of age and younger.

**Conclusion:** The above findings indicate that asymptomatic carriage of *P. falciparum* in children living in a hyperendemic area occurs independently of IgG but is associated with a balanced inflammatory cytokine ratio.

## Introduction

Malaria is still a significant global health problem with an estimated 249 million cases and 608 000 deaths in 2022 [1]. Although mortality rates from malaria have decreased considerably since the turn of the century [2] this has been paralleled by high increases in proportions of asymptomatic infections, known to contribute significantly to persistent malaria transmission in most endemic countries [3,4]. Asymptomatic infections can persist for weeks to years [5] and untreated *P. falciparum* infections have been shown persist on average for 6 months, with fluctuations in parasite density over the course of an infection [6,7]. The maintenance of chronic asymptomatic malaria will depend on the host immune response to the blood stage of *Plasmodium* [3]. Anti-parasite immunity directly suppress parasite growth resulting in low infection densities (below the pyrogenic threshold [3,8]) whilst anti-disease immunity modulates inflammation-mediated clinical manifestations of the infection.

Older children and adults have more cumulative exposure to parasite variants circulating within the community, developing adaptive responses which mediate both anti-parasite and anti-disease immunity [9,10]. However, asymptomatic infections have been documented in younger children < 5 years who are too young to have developed robust adaptive immunity [11]. This suggests that asymptomatic malaria may be differentially controlled by innate and adaptive immune responses depending on age.

Anti-parasite immunity which is acquired with age or exposure [12,13], is mediated mainly by antibodies which control parasite replication and mediate clearance of *Plasmodium*-infected red blood cells (iRBCs) [14–16]. Suppression of inflammation associated with iRBCs depends on the balance between anti- and pro-inflammatory cytokine responses [17,18] and it has been hypothesized that asymptomatic infections are associated with a shift in host immune responses towards an anti-inflammatory profile after multiple exposures to malaria [19]. This could occur via a decrease in levels of pro-inflammatory immune mediators such as IFN-γ, TNF-α, or an increase in anti-inflammatory mediators such as IL-10 or TGF-β [18,20–22].

The relative contributions of anti-parasite and anti-disease immunity to the maintenance of persistent asymptomatic *Plasmodium* infections in younger individuals and older individuals remain unclear. To determine the immunological associations of chronic asymptomatic malaria in younger (≤20 years) and older (>20 years) individuals we have undertaken a longitudinal study of highly exposed individuals of all ages with asymptomatic malaria over a 3-month period where we monitored the stability of IgG and cytokine responses. We also examined these responses upon clearance of parasites to ascertain which responses depended on the presence of circulating parasites. Our study demonstrates a lack of correlation of iRBC density and IgG responses in individuals ≤20 years old, with an association of parasite carriage with IL-10 and an association of asymptomatic status with a balanced ratio of IL-10:TNF-α and IL-10: IFN-γ.

## Material and methods

### Ethics statement

The study protocol was approved by the Cameroon National Ethics Committee for Human Health Research (Ethical clearance N°: 2018/09/1104/CE/CNERSH/SP) and the Ministry of Public Health. Written informed consent was obtained from each participant or parent/guardian of minor participants (< 19 years old) prior to enrolment into the study. Assent was obtained from all children older than 12 years of age.

### Study area and population

The study population consisted of individuals of both genders aged at least two years of age and permanently living in one of the five selected villages of the Esse health district, Mefou-et-Afamba Division, Centre Region of Cameroon where malaria transmission is perennial, stable and hyper-endemic with *P. falciparum* as the predominant *Plasmodium* species. The study area has been described elsewhere [23–25]. The inclusion criteria were: i) obtain the consent of each older participant or parent or legal guardian for the children, ii) living permanently in the study area, iii) not pregnant.

### Study design and participants

This was a longitudinal household-based study that involved 353 individuals. Participants were considered asymptomatic for malaria if positive for *P. falciparum* using multiplex nested PCR in the absence of fever at least forty-eight hours prior to enrolment. Being febrile was assessed based on axillary temperature ≥ 37.5 °C or based on the history of fever as independently confirmed by another household member. All symptomatic individuals (fever + rapid diagnostic test (RDT) +) were treated with ACT-based drugs following recommended national guidelines. Participants with asymptomatic carriage of *Plasmodium* parasites were monitored weekly until they either developed a fever, or until the end of the follow up period at week 16 post-enrolment. Participant follow-up comprised of weekly interviews regarding the occurrence of fever during the past week, measurement of axillary temperatures on each visit, and laboratory testing for malaria at weeks 1, 4, 11 and 16 (PCR and microscopy). All symptomatic cases occurring during the follow-up period were treated with ACT as recommended and excluded from further follow-up. At week 11 post-enrolment all participants with persisting asymptomatic parasitemia were treated as recommended, and monitored for five additional weeks to assess the effect of ACT-based antimalarial treatment on selected host immune responses. Participants with persistent asymptomatic parasitemia 10 weeks following enrolment were considered as persistent asymptomatic malaria whereas those who developed malaria-associated symptoms before week 11 post-enrolment were considered as developing uncomplicated malaria.

### Blood sample collection, malaria diagnosis and parasite genotyping

Approximately 3 ml of venous blood was collected in EDTA-coated tubes and used for laboratory testing for malaria and blood hemoglobin levels as well as to obtain plasma for immunological analysis. Plasma was separated from the red blood cell (RBC) pellet by centrifugation (3500 RPM, 5 min) and stored at −80 °C until use. Blood hemoglobin levels were assessed using a portable Mission Hb haemoglobinometer (ACON Laboratories, USA).

Parasite density (parasites/µl) was determined by microscopic examination of Giemsa-stained thick blood smears. Parasite density was determined based on the number of parasites per at least 200 WBC count on a thick film, assuming a total white blood cell count of 8000 cells/μL of whole blood. A blood slide was considered positive when a concordant result was obtained by at least two independent microscopists and when a discrepancy occurred, a third microscopist was used for confirmation. A slide was considered negative if no parasites were detected after a count of 500 white blood cells.

Genomic DNA was extracted from dried blood spots using the Chelex method [26], and used to screen for *Plasmodium* infections by multiplex nested PCR targeting the 18S small subunit rRNA (ssrRNA) genes as previously described [24]. Genotyping of *msp2* gene was performed by nested PCR on *P. falciparum* PCR positive samples as previously described [24,27].

### Case definitions for the study

Participants were defined as asymptomatic if positive for malaria parasites by PCR in the absence of fever at least 48 hours prior to enrolment. Febrile malaria was determined based on RDT, light microscopy or PCR positivity for malaria parasites and axillary temperature ≥ 37.5°C or a history of fever in the past 48 hours. Those developing uncomplicated malaria were defined as asymptomatic individuals who had developed fever associated with *Plasmodium* infection during the 10-week infection follow-up period, whereas the persistent asymptomatic malaria classification represented the asymptomatic individuals who remained asymptomatic until anti-malarial treatment at the end of the infection follow-up period.

### Anti-*Plasmodium* antibody levels and avidity assessment

Plasma IgG antibody levels and avidity were determined by indirect ELISA method using *P*. *falciparum* Merozoite extract (ME), schizont extract (SE) or the recombinant proteins erythrocyte binding antigen-175 (EBA-175) and Merozoite Surface Protein-1_19_ (MSP1_19_) as coating antigens. ME and SE were isolated from laboratory maintained *P. falciparum* 3D7 parasites as described previously [28], and used for protein extraction by freeze-thaw fractionation [23,29]. The recombinant protein EBA-175 (F3 region) was obtained from BEI Resources (MRA-1162), whereas *P. falciparum* MSP1_19_ was produced using a baculovirus-insect cell expression system [30].

96-well microplates (F96 CERT-Maxisorp) were coated with each antigen (2 mg/ml of ME and SE, 0.20 mg/ml of EBA-175 and 0.5 mg/ml of MSP-1_19_) in 0.1 M bicarbonate buffer (for ME, SE and EBA-175) or PBS (for MSP-1_19_) and incubated overnight at 4°C. Plasma samples were diluted (1/250) in PBS-T/1%BSA and used to measure anti-*Plasmodium* IgG as previously described [23,31]. Plasma from non-exposed European blood donors (10 samples) were used as naive controls and a pool of IgG fractions from adults living in high malaria endemic areas was used as positive controls. Results were expressed as optical-density ratios (OD sample/mean OD naive control). For avidity testing, 100 µL of 8M urea in wash buffer (treated wells) or 100 µL of wash buffer (untreated wells) were added for 15 minutes at room temperature to the respective wells following primary antibody binding as previously described [32]. Avidity indices (AI) were calculated as proportion of bound antibodies following urea treatment (AI = [mean OD of treated wells /mean OD of untreated wells] × 100). Samples with OD ratios > 2, corresponding to mean OD naive control + 3SD were considered as seropositive. In addition, only seropositive samples were used for avidity testing.

### Growth inhibition assay (GIA)

GIA was performed using plasma from endemic and non-endemic individuals as described by Duncan *et al.* [33]. Briefly, plasma samples were heat inactivated at 56 ° C for 20 min and a 1:20 ratio of 50% RBC was added to each sample and incubated at room temperature for 1 h to remove the RBC-reactive antibody. After spinning for 2 min at 13,000 × *g* to pellet the RBCs, the supernatant was transferred to a 96-well V-bottom plate. Synchronized late trophozoite/early schizont (30-40 h) stages of 3D7 *P. falciparum* at 0.5% parasitemia were incubated with plasma (1:10 ratio) for 48 h at 37 °C. The GIA harvest (parasite growth) was carried out by SYBR green-I DNA quantification assay [34]. Parasite growth inhibition (GI) activities of plasma antibodies were presented as: GI indices (%) = 100 × [1 − ((OD sample − OD RBC) / (OD parasitized RBC − OD RBC))].

### Determination of plasma cytokine levels

Plasma concentrations of 38 immune mediators including interleukin (IL)-1α, IL-1β, IL-1RA, IL-2 IL-3, IL-4, IL-5, IL-6, IL-7, IL-8, IL-9, IL-10, IL-12p40, IL-p70, IL-13, IL-15, IL-17A, interferon (IFN)-α2, IFN-γ, tumor necrosis factor (TNF)-α, lymphotoxin-α (LT-α), IFN-γ induced protein (IP-10), monocyte chemoattractant protein (MCP)-1, MCP-3, macrophage inflammatory protein (MIP)-1α, MIP-1β, Eotaxin, Fractalkine, human growth-regulated oncogene (GRO), Macrophage-derived chemokine (MDC), granulocyte-macrophage colony-stimulating factor (GM-CSF), granulocyte colony-stimulating factor (G-CSF), sCD40L, FMS-like tyrosine kinase 3 ligand (Flt-3L), epidermal growth factor (EGF), fibroblast growth factor 2 (FGF-2), transforming growth factor alpha (TGF-α) and vascular endothelial growth factor (VEGF) were determined by Luminex-based method using the Human Cytokine/Chemokine Magnetic Bead Panel MILLIPLEX MAP Kit (Cat. No. HCYTMAG-60K-PX38, EMD Millipore Corporation) according to the manufacturer’s instructions. Briefly, 25 µL of premixed magnetic capture beads were incubated with equal volumes of plasma or diluted standard in a 96-well microplate placed on a horizontal plate shaker for 2 hours. After washing using a magnetic plate, plates were incubated with 25 µL/well of biotinylated detection antibodies for 1 hour at room temperature followed by 25 µL of streptavidin-PE in each well for 30 minutes at room temperature. Median fluorescence intensity was read on a Luminex MAGPIX Analyser (XMAP Technology) equipped with xPONENT software version 4.2. The relative concentration of each cytokine (pg/ml) in each sample was determined based on the automatically generated standard curve for each analyte. Analyte amounts less than the limit of detection for each cytokine were attributed the value of detection limit.

### Statistical analysis

Statistical analyses were performed using RStudio version 4.1.1 or version 4.2.2 [35] and Graphpad Prism version 8.4.3 (Graphpad software, San Diego, California USA). Differences in antibody and cytokine distribution between two or more groups were determined using Wilcoxon signed rank test and Kruskal-Wallis test, respectively. Spearman rank correlation tests or linear regression were used to determine the correlation between quantitative variables. Differences in categorical variables were determined by chi-square tests. P-values < 0.05 were considered statistically significant. Factor analysis of mixed data (FAMD) with FactoMineR and factoextra packages was used to assess the association between cytokines levels and age and circulating parasite loads. The complexHeatmap and ggpubr packages were used to draw the heatmaps and the line plots, respectively. Given that the aim of this study was to determine the host immunological determinants in the maintenance of the asymptomatic infections, among the 353 individuals included, only participants with complete information on the study outcome during the longitudinal follow-up period were included in the analysis (n= 211). Participants who missed at least one of the follow-up points (the predominant reason being farming far away from the enrolment site) were not included in our study. Three samples (2 symptomatic and 1 asymptomatic) were excluded because no cytokines were detected in the plasma. For analysis of cytokines at the initial time point of our study with respect to correlation with persistent asymptomatic malaria, we employed a mixed linear model using the LME4 package [36]. Cytokine level was used as the response with conversion status as the fixed-effect of the model. We used sex and age as random effects in the model and ran the model separately for children ≤20 years old, adults >20 years old and all participants, regardless of age. To obtain p-value estimates from the t-values we used the lmerTest package [37]. P values were corrected for multiple comparisons using the Sattherthwaite correction. For analysis, parasite densities were separated into three groups as follows, 1. submicroscopic parasitemia, 2. low levels of microscopic parasitemia (parasitemia between 37.2 parasites/μl and 413.1 parasites/μl) and 3. high levels of parasitemia (parasitemia> 413.1 parasites/μl, representing the median parasitemia in the study population).

## Results

### Asymptomatic malaria prevalence in the study area

We carried out immunological analysis of a subset of 211 individuals from a previously reported study of 353 individuals screened for malaria using PCR, where 328 (92.9%) were positive for *Plasmodium falciparum* infection, of which 266 (81.1%) were asymptomatic and 62 (18.9%) were symptomatic [24]. This subset had complete sampling from the follow-up study, and included 142 individuals with asymptomatic infection, 61 symptomatic individuals and 8 non-infected individuals. At baseline when these individuals were first sampled, parasite densities were highly variable between the symptomatic and asymptomatic groups, and no significant difference was observed in the median parasite densities of the two groups at the initial time point of the study (**Table 1**). Age, sex, and hemoglobin levels were also not different across the study groups at this initial time point (**Table 1**).

**Table 1.** Characteristics of the study population. The data presented in this table is from 211 individuals from our study at baseline. The subset of 142 asymptomatic individuals included in this table were those individuals with complete data from the follow-up study to 11 weeks and does not include baseline data from individuals who dropped out of the study. P-value was determined using Wilcoxon signed-rank test and Kruskal-Wallis test when comparing two or three groups, respectively. The median parasitemia presented in this table is only for the infected individuals with parasite densities at the detection limit of light microscopy. The sex ratio between groups were compared using chi-square test. Hb: hemoglobin. M: male. F: female.

| Variables | Non detectable infection (n= 8) |  | Asymptomatic (n= 142) |  | Symptomatic (n= 61) |  | P value |  |
| --- | --- | --- | --- | --- | --- | --- | --- | --- |
| Age groups | ≤ 20 | > 20 | ≤ 20 | > 20 | ≤ 20 | > 20 | ≤ 20 | > 20 |
| n | 5 | 3 | 94 | 48 | 42 | 19 |  |  |
| Sex ratio (M:F) | 0.67 (2:3) | 50 (1:2) | 0.82 (42:51) | 41.2 (14:34) | 1.1 (22:20) | 58.3 (7:12) | 0.7114 | 0.8280 |
| Median age, years (range) | 14 (4-20) | 68 (50-68) | 9 (2-19) | 50 (22-86) | 9 (2-20) | 28 (21-64) | 0.2193 | 0.0028 |
| Median Hb, g/dl (range) | 11.8 (10.7-12.7) | 112.5 (11.9-14) | 12 (4.9-14.3) | 13.1(10.9-16) | 11.1 (6.8-14) | 12.8 (9.4-16.8) | 0.1866 | 0.9833 |
| % of submicroscopic % (n) | - |  | 22.3 (21) | 60.4 (29) | 9.5 (4) | 57.9 (11) | 0.0746 | 0.8495 |
| Geometric mean parasitemia, parasites/μl (range) | - |  | 575.7 (37.4-68190.5) | 113.5 (38.3-660.2) | 764.1 (38.1-240457.1) | 183.3(37.2-5058.8) | 0.5139 | 0.6101 |

### Anti-parasite antibodies were inversely correlated with parasitemia but were not associated with asymptomatic parasite carriage

Consistent with the hypothesis that host immunity increases with repeated exposure to malaria parasites, baseline parasite densities above the detection limit of microscopy decreased significantly with age (r= −0.49, P< 0.0001) (**Figure 1A**) but only in individuals aged 20 years and younger (r= −0.39, P< 0.0001) (**Figure 1B**). Of the 142 participants who completed the follow-up study as designed, 78 (54.9%) remained asymptomatic during the 10-weeks follow-up period, whereas 63 (44.4%) developed malaria-associated fevers and 1 (0.7%) resolved the infection in the absence of any known antimalarial treatment. Based on the parasitemia distribution of those with microscopic *Plasmodium* infections at baseline the study population was divided into two groups aged ≤ 20 years and > 20 years old for further analyses. The proportion of submicroscopic infections was significantly higher (P<0.0001) in the older age group (age >20 years) at the baseline time point than in the younger age group (**Figure 1C**). At the baseline time point, median IgG levels against *P. falciparum* merozoite (ME) and schizont (SE) crude extracts were significantly higher in the >20-year age group than in the younger age group (P< 0.0001) (**Figure 2A**). This was mirrored by IgG reactive to *P. falciparum* recombinant proteins EBA-175 (P= 0.0250) and MSP-1_19_ (P< 0.0001) (**Figure 2A**). Additionally, In the younger age group IgG levels against merozoite (r= 0.22, P= 0.0077) and schizont (r= 0.29, P= 0.0004) crude extracts as well as against MSP-1_19_ (r= 0.15, P= 0.0764) but not EBA-175 (r= 0.01, P= 0.8525) were positively correlated with age at the baseline time point (**Figure 2B**). A negative correlation was observed between IgG levels to *P. falciparum* merozoite and schizont extracts at baseline and parasite density measured at the same time point (**Supplementary Figure 1**). Indeed, the multilevel linear regression model adjusted for village and household levels, using IgG levels at the baseline and age as predictors, showed that only age was the independent predictor of *Plasmodium* parasite densities in the study population (coef= −0.02, P= 0.0004 for age; coef= 0.01, P= 0.9170 for ME; coef= −0.19, P = 0.1529 for SE; coef= −0.02, P= 0.4746 for EBA-175; coef= 0.02, P= 0.5362 for MSP-1_19_).

**Figure 1.**
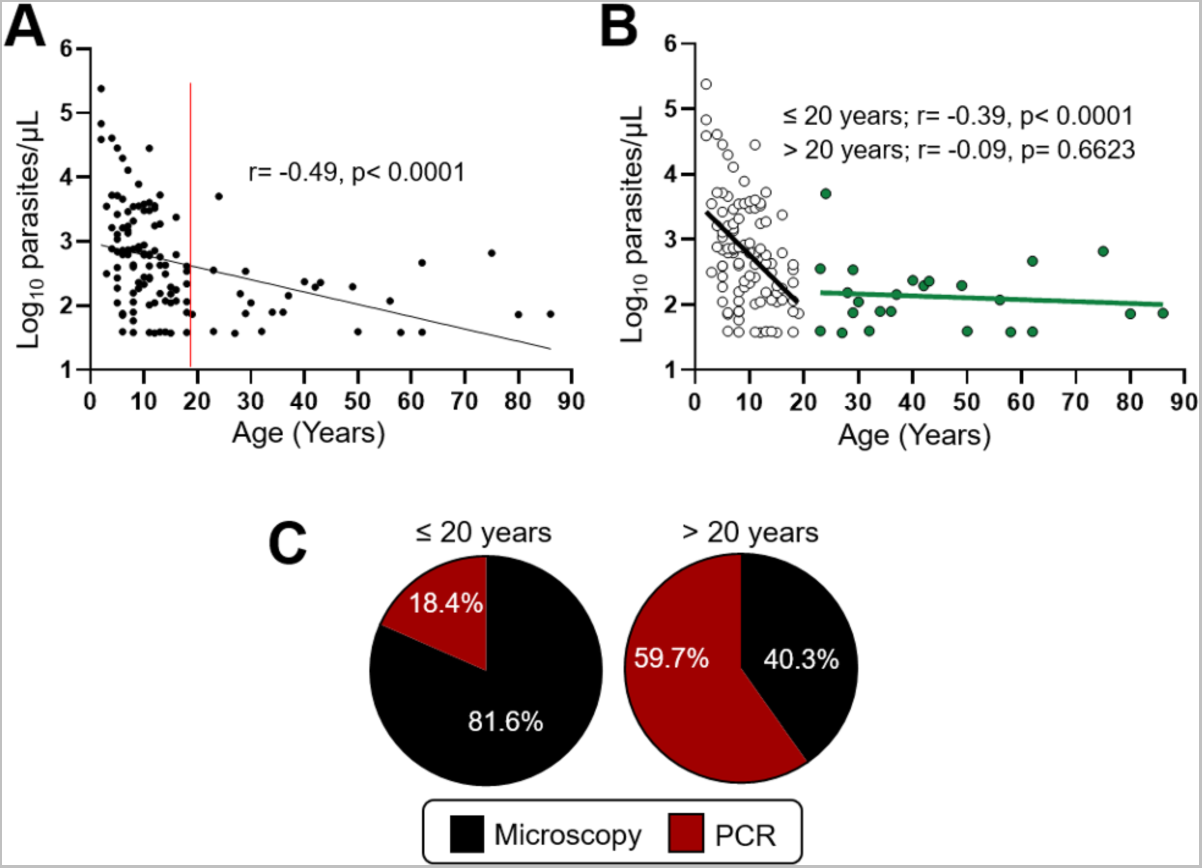
Older individuals (>20 years) had a better control of parasite densities. **A**) Scatterplot showing a correlation between baseline parasite densities and age in all the infected individuals with parasitemia at the detection limit of light microscopy (n= 138). Spearman rank correlation is presented as the best fit line and the coefficient (r), as well as the p-value (p), are shown. The red straight line was used to divide the study population into two groups aged ≤ 20 years and > 20 years old for further analyses. **B**) Scatterplot of correlation between baseline parasitemia detected by microscopy and age in different age groups (≤ 20 years and > 20 years) at the different time points of our study. Spearman rank correlation is presented as the best fit line and the coefficient (r), as well as the p-value (p), are shown for each age group. **C**) The pie chart shows the proportion of sub-microscopic infections (PCR+/microscopy-) versus microscopic (microscopy+) infections in each age group. PCR: Polymerase Chain Reaction.

**Figure 2.**
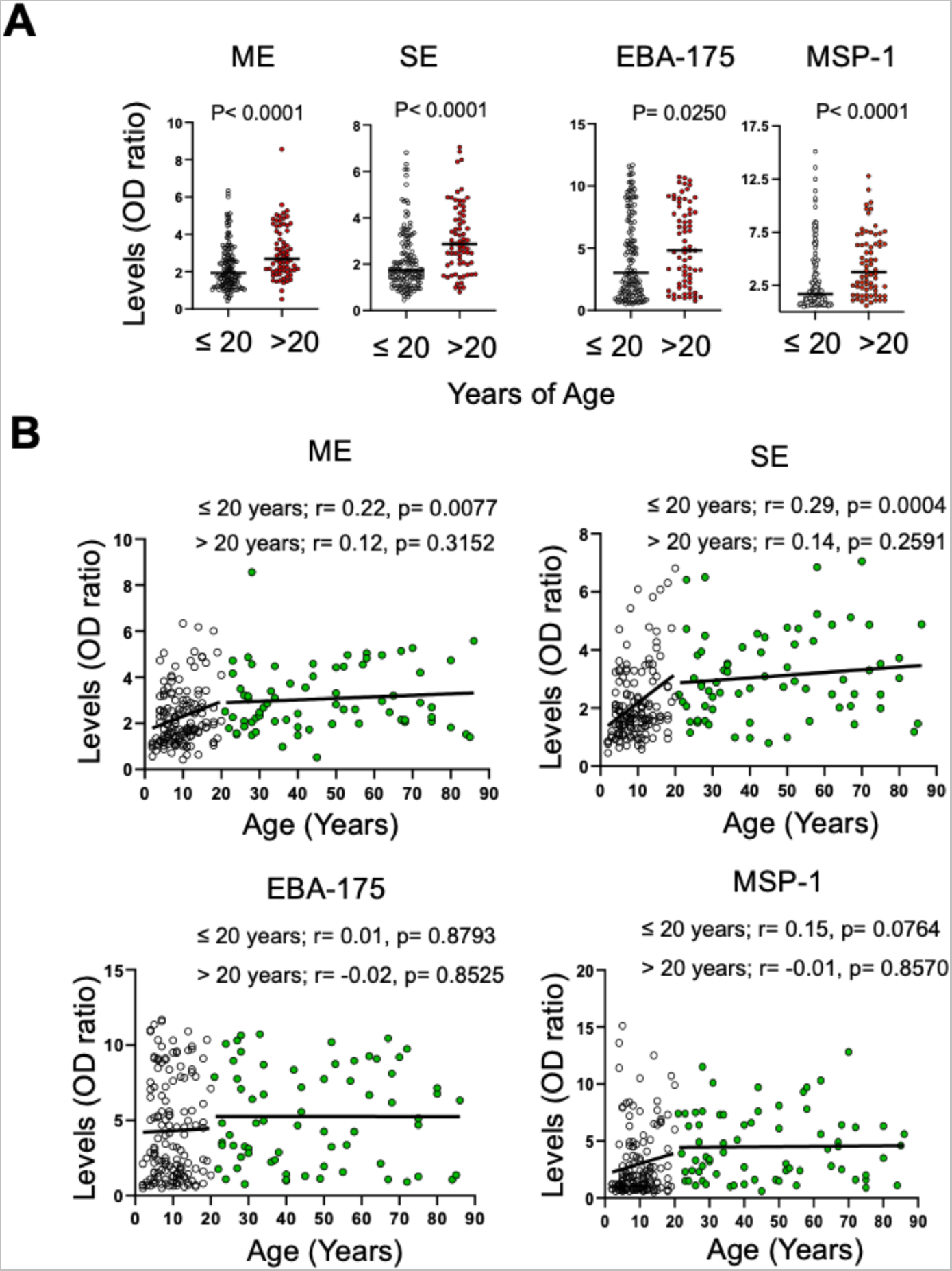
Individuals >20 years old had more anti-parasite IgG at baseline. **A**) Data presented in the scatterplot represent baseline IgG antibody levels against different *P. falciparum* antigens (ME, SE, EBA-175 and MSP-1_19_) within age groups. Median levels between age groups were compared by Wilcoxon signed-rank test and the p-values are shown on the graph. **B**) The scatterplots on B show the correlation between IgG levels against different *P. falciparum* antigens (ME, SE, EBA-175 and MSP-1_19_) and age in different age groups (≤ 20 years and > 20 years). Spearman rank correlation is presented as the best fit line and the coefficient (r), as well as the p-values (p), are shown for each age group. SE: Schizont extract. ME: Merozoite extract. EBA-175: Erythrocyte binding antigen-175. MSP-1_19_: Merozoite surface protein-1_19_. OD: optical density.

Median IgG levels were not significantly different between participants with sub-microscopic parasitemia and those with microscopic parasites at baseline (**Supplementary Figure 2A**), and no significant difference was observed in the *in vitro* growth inhibition potential of inactivated plasma from submicroscopic parasite carriers and that of microscopic carriers (**Supplementary Figure 2B**). Both IgG antibody avidity (**Supplementary Figure 3A&B**) and growth inhibition potentials (**Supplementary Figure 2B**) were similar in the two groups (P> 0.05). Together, these findings suggest that anti-parasite antibodies may control parasite density levels by mechanisms other than direct inhibition of parasite growth *in vivo*.

Similarly, no significant difference was observed in malarial IgG antibody levels and avidity between participants with symptomatic or asymptomatic parasitemia (**Supplementary Figure 4A**), and no significant difference was seen in the blood parasitemia at the detection limit of microscopy (Supplemental Figure 4B; **Table 1**). However, the *in vitro* growth inhibition potential of plasma from asymptomatic malaria was greater than that of symptomatic individuals (P= 0.0245) despite no significant differences in the parasitemia amongst those with parasite levels that were detectable by microscopy (**Supplementary Figure 4B**). This may be a reflection of higher percentage of asymptomatic children with submicroscopic parasitemias (77.7%) compared to symptomatic children (90.5%) (**Supplementary Figure 4C**). Taken together, these data suggest that humoral responses other than IgG responses to the tested parasite antigens contribute to the maintenance of asymptomatic parasitemia in the study area.

### Persistence of fluctuating asymptomatic parasitemia occurred in highly exposed individuals without alteration in IgG levels

Of the 142 participants who completed the follow-up study as designed, 72 (50.7%) remained asymptomatic during the 10-weeks follow-up period, whereas 69 (48.6%) developed malaria-associated fevers and 1 (0.7%) resolved the infection in the absence of any known antimalarial treatment. Surprisingly, the propensity to convert was not associated with age (**Figure 3A**). Of the 72 participants who maintained chronic asymptomatic *P. falciparum* carriage, the proportion of submicroscopic infections remained similar in both age groups with most participants becoming microscopy positive on week 10 regardless of age (**Figure 3B**). Parasite densities fluctuated widely over the follow-up period from ultra-low parasitemia (sub-microscopic) to high parasitemia (**Figure 3B**), possibly due to variation in the infecting clones from new infections as this is a high transmission area (**Figure 3C**). However, no changes in IgG levels (**Figure 3D**) against all antigens tested were observed during persistent asymptomatic infections indicating that persistent asymptomatic parasitemia does not affect anti-*Plasmodium* IgG levels over this short 3-month time period. These data suggests that asymptomatic carriage of *P. falciparum* occurs independently of circulating parasite numbers, even within highly-exposed individuals.

**Figure 3.**
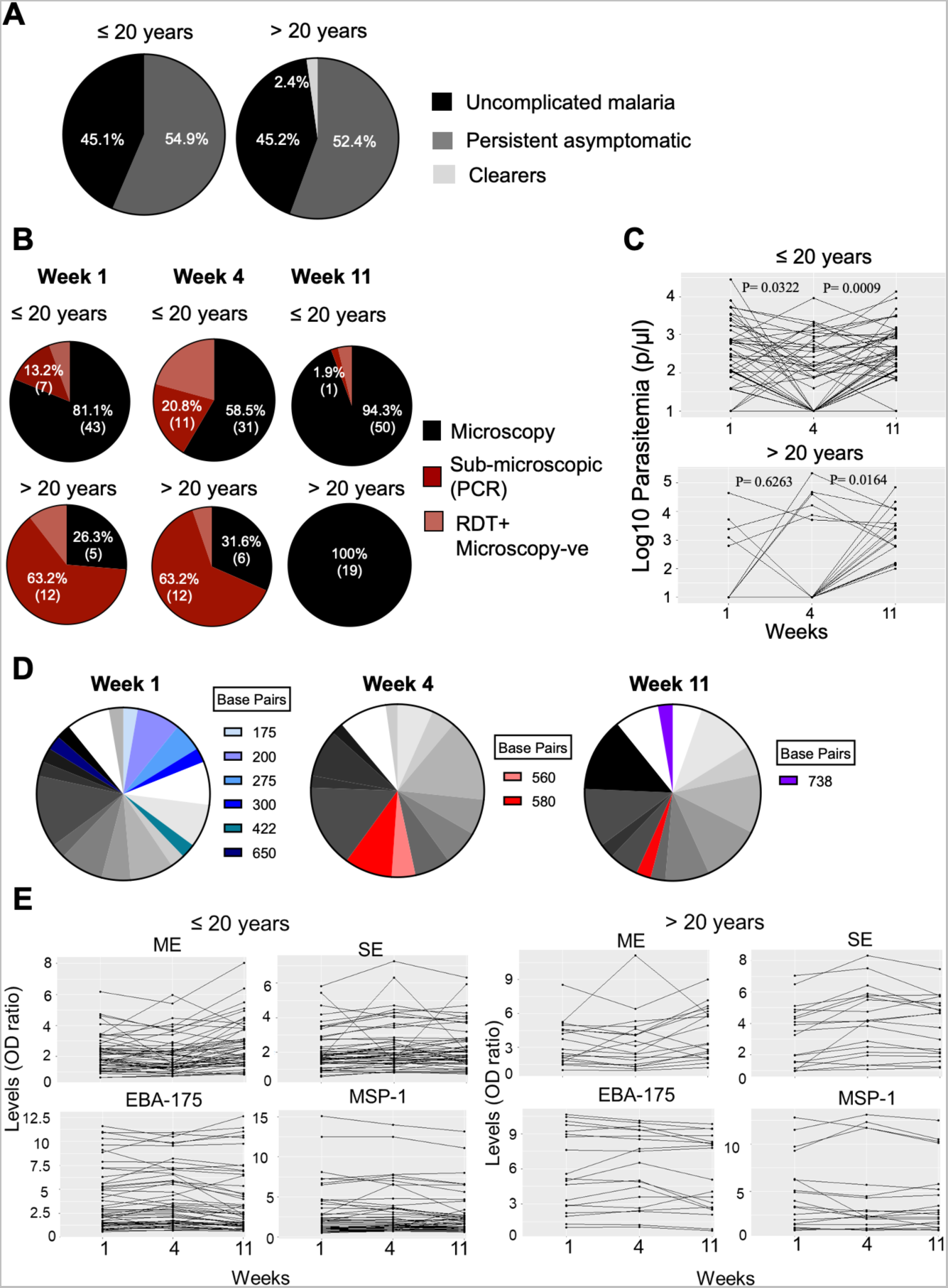
Asymptomatic malaria persisted in some individuals despite changes in the circulating parasite densities and population over time, but did not affect the antibody levels. **A**) The pie charts show the proportion of individuals with initial asymptomatic infection (cross-sectional) who resolved the infection (clearers), developed malaria-associated fever (uncomplicated malaria) or remained asymptomatic (persistent asymptomatic) over the 10-week follow-up period, according to age groups. **B**) Panel B displays the proportion of sub-microscopic/microscopic parasitemia in persistent asymptomatically infected individuals at three different time points (weeks 1, 4 and 11) according to age groups. Individuals who were RDT positive but without detectable micoroscopic parasitemia was detected are shown in pink. Of those who had microscopic parasitemia, the line plots parasitemia across the 3 time points (**C**). To generate the line plots, the value 10 was assigned to submicroscopic parasitemia. The P values on the line plots refer to the comparison of the median parasitemia between two time points using the Wilcoxon test. **D**) Pie charts show the proportion of msp2 alleles (representing the clones) determined by nested PCR in individuals with persistent asymptomatic infection per week. Numbers represent the band sizes obtained. Blue slices represent the alleles only present at week 1. Red slices represent the alleles which appear at week4. Green slices represent the allele only present at week 11. Grey slices represent the alleles that were present at all three sampling time points (bands of 375, 400, 475, 500, 520, 534, 540, 600, 620, 633. 680, 700, 800 bp). **E**) Data presented in the line plots represent the profile of IgG anti-*Plasmodium* antibody levels in each chronic asymptomatically infected individuals at three different time points (week 1, 4 and 11) in the two age groups. SE: Schizont extract. ME: Merozoite extract. EBA-175: Erythrocyte binding antigen-175. MSP-1_19_: Merozoite surface protein-1_19_. OD: optical density.

### Parasite density was associated with inflammatory responses amongst younger individuals with asymptomatic malaria

Given that *Plasmodium* parasites are associated with induction of inflammatory responses we measured the cytokine profiles of the 72 participants who had persistent asymptomatic carriage of *P. falciparum* across the course of our study. There was no particular cytokine signature that associated with asymptomatic carriage of *P. falciparum* in our study participants over the 11-week follow-up period (**Figure 4A**). Using a factor analysis of mixed data (FAMD) on cytokine data along with age and circulating parasite density, we found that levels of IL-10, TNF-α, the ratio of IL-10:IFN-γ and MCP-1, along with age and parasite density explained most of the variation in the peripheral cytokine response in asymptomatic individuals (**Figure 4B**). Multivariable analysis using age and parasitemia as predictors showed that age independently predicted the levels of TNF-α (coef= −0.31, P< 0.0001) peaking in younger study participants (**Figure 4C**). This is likely a reflection of circulating parasite densities which were higher in the younger group (**Figure 1**) and were also independently predictive of TNF-α levels (coef= 0.09, P< 0.0001) (**Figure 4D**). Circulating parasite density also independently predicted the levels of IL-10 (coef= 0.21, P< 0.0001) and MCP-1 (coef= 0.06, P= 0.01) with a significant increase in those who had high levels of parasitemia (parasitemia> 413.1 parasites/µl) relative to those who had low levels of microscopic parasitemia (parasitemia between 37.2 parasites/µl and 413.1 parasites/µl) (**Figure 4D**). This indicates that during asymptomatic carriage of *P. falciparum* circulating parasites simultaneously induce anti-inflammatory cytokine production to counteract the inflammatory effects of TNF-α. To determine which cytokines were dependent on parasite carriage we treated participants at week 11 of the study with ACT. This resulted in a significant decline in the IL-10 levels, but not TNF-α or MCP-1 levels, in the younger age group (**Figure 5**). Thus, the levels of IL-10 measured is a reflection of parasite carriage in the younger age group.

**Figure 4.**
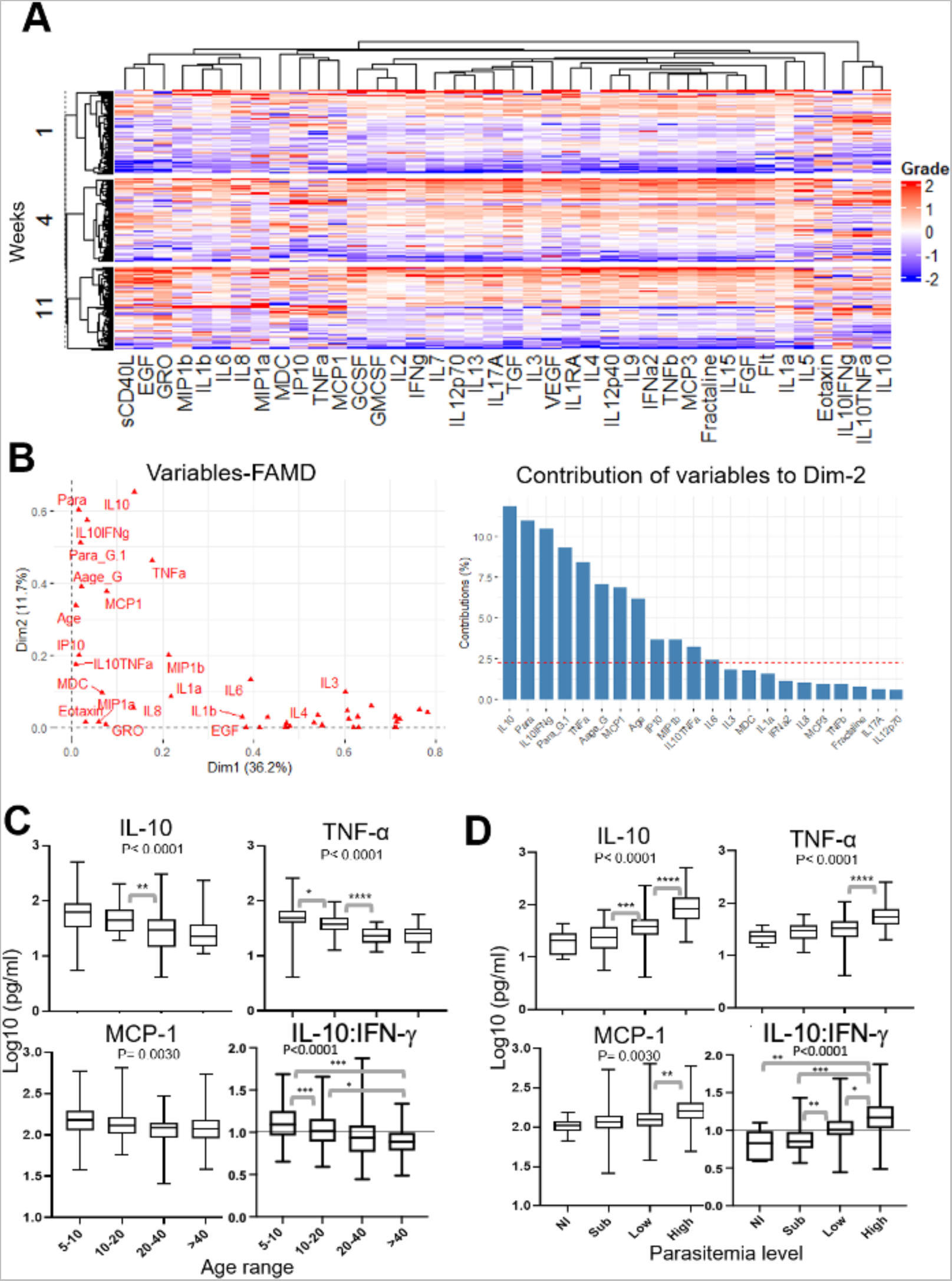
Cytokine responses were blunted with increased age and correlated with parasite densities at baseline. **A**) Hierarchical clustering of the standardized least-square means of 38 differentially expressed cytokines/chemokines/growth factors across the three sampling time points in participants with persistent asymptomatic infection (n= 72). **B**) Principal component of cytokines, age and parasitemia from FAMD (left graph) analysis. The right graph of B shows the contribution of each variable on the principal component 2 (dimension 2). **C** and **D)** box plots representing interquartile ranges (25th, median and 75th percentile, whisker percentile (1st and 99th)) of the levels (Log10 pg/ml) of some main regulatory cytokines measured by Luminex according to age and parasite level groups at the baseline, respectively. Median levels of cytokines between two or more groups were compared by Wilcoxon rank-sum test and Kruskal-Wallis test, respectively. In Panel A, pairwise comparisons on neighboring groups were performed only when multiple comparisons were significant at p< 0.05. Parasite densities were groups in three groups as follows: submicroscopic parasitemia, low levels of microscopic parasitemia (parasitemia ≤ 413.1 parasites/μl, representing the median parasitemia in the study population) and high levels of parasitemia (parasitemia> 413.1 parasites/μl). FAMD: Factor analysis of mixed data. *p< 0.05; **p< 0.001, ***p< 0.0001, ****p< 0.00001.

**Figure 5.**
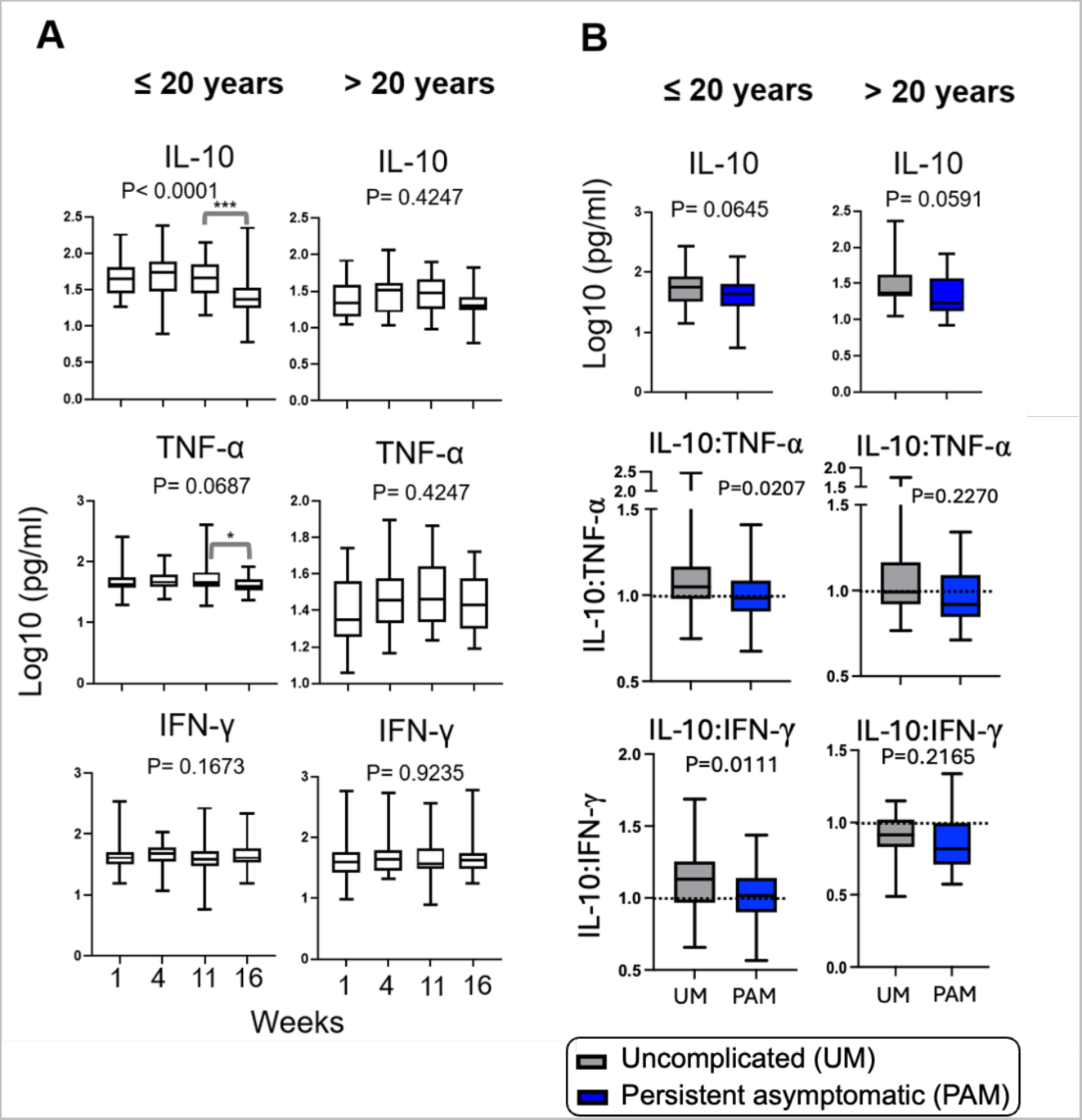
IL-10 and TNF-α levels were a reflection of parasite carriage, and the balance of inflammatory cytokines at baseline was associated with the maintenance of asymptomatic infections in the younger age, ≤ 20 years. **A**) Box plots representing interquartile ranges (25th, median and 75th percentile, whisker percentile (1st and 99th)) of cytokine levels (Log_10_ pg/ml) during persistent asymptomatic infections (week 1, 4 and 11) and after anti-malarial treatment (week 16). **B**) Box plots representing interquartile ranges (25th, median and 75th percentile, whisker percentile (1st and 99th)) of cytokine levels (Log_10_ pg/ml) at the baseline between in those who developed uncomplicated malaria (UM; grey) and those who went on to display persistent asymptomatic malaria throughout the course of the study (PAM; blue). Analyses were performed including values from individuals with submicroscopic and microscopic parasitemia. Median cytokines levels between two or more groups were compared by Wilcoxon rank-sum test and Kruskal-Wallis test, respectively and significant between group differences indicated by the P value on the plot. In Panel A, pairwise comparisons on neighboring groups were performed only when multiple comparisons between the 4 groups were significant and are shown by the grey brackets. *p< 0.05 and ***p< 0.0001.

### Maintenance of asymptomatic carriage of *P. falciparum* is associated with a more balanced inflammatory cytokine ratio

There was no particular cytokine signature of asymptomatic individuals relative to symptomatic individuals in our study (**Figure 6A**). However, given that anti-inflammatory cytokines may be important in down-regulating the pathogenic inflammatory response in malaria, we tested the hypothesis that asymptomatic infection would be associated with higher levels of IL-10. To our surprise, at the initial time point of our study there was a non-significant trend towards higher levels of IL-10 in the symptomatic compared to asymptomatic participants (**Figure 6B**). This may be a reflection of higher inflammation levels in symptomatic individuals as other pro-inflammatory markers (IL-6 and MCP-1) were significantly higher in individuals with symptomatic malaria compared to those with asymptomatic *P. falciparum* infections in the younger age group (**Figure 6B**). There was an IL-10-skewed ratio of IL-10:TNF-α and IL-10:IFN-γ in the symptomatic group compared to the asymptomatic group in both age groups (**Figure 6C**). This is in agreement with previously published data in individuals with uncomplicated malaria and the IL-10 skewed ratio of IL-10 to TNF-α that can be measured in those with severe symptoms of malaria [38,39]. Multivariable logistic regression analysis showed that the IL-10:TNF-α ratio and IL-1RA levels were independently associated with developing symptoms during *P. falciparum* infection in this study (**Supplementary Table S1**).

**Figure 6.**
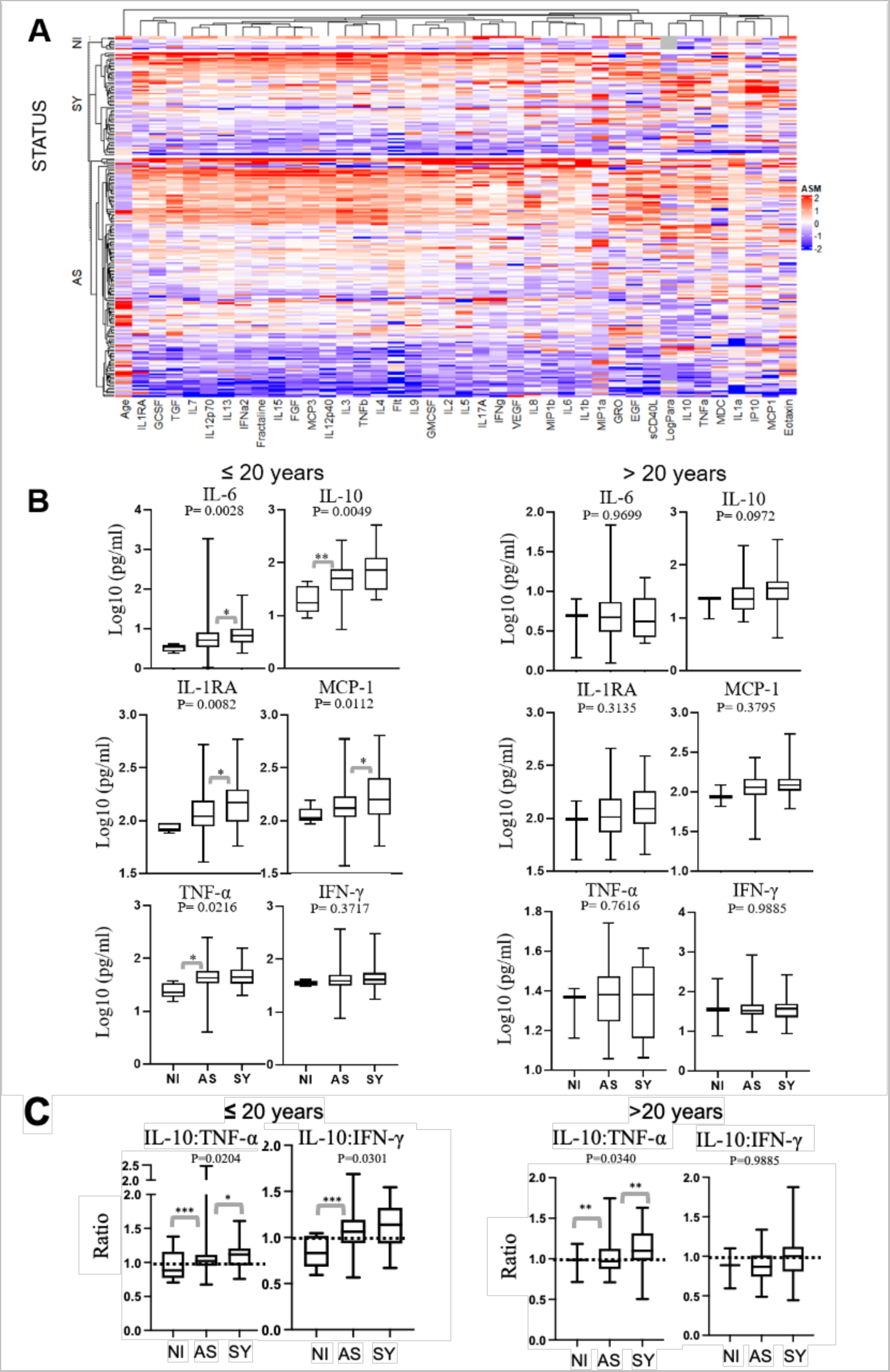
IL-10 at baseline was associated with clinical malaria in the younger age group (≤ 20 years). **A**) Hierarchical clustering (heatmap) of the standardized least-square means of 38 differentially expressed cytokines/chemokines/growth factors by infection status at the baseline of this study. **B** and **C** box plots represent interquartile ranges (25th, median and 75th percentile, whisker percentile (1st and 99th)) of the baseline cytokine levels (Log_10_ pg/ml) measured by Luminex according to infection status. Analyses were performed including both individuals submicroscopic and microscopic parasitemia. Median levels of cytokines between two or three groups were compared by Wilcoxon rank-sum test and Kruskal-Wallis test, respectively. In Panel B and C, pairwise comparisons were performed only when multiple comparisons were significant. P values refer to the Kruskal-Wallis test comparison between the three groups. NI: Non-infected. AS: Asymptomatic. SY: Symptomatic. *p< 0.05; **p< 0.001; ***p< 0.0001.

To further test the hypothesis that asymptomatic carriage of *P. falciparum* may be related to IL-10 production, we compared the cytokine ratios in the younger group who converted to symptomatic malaria throughout our study with those who remained asymptomatic. At the initial time point of the study there was a trend towards increased IL-10 levels in those who subsequently went on to develop uncomplicated malaria compared to those who experienced persistent asymptomatic malaria in both age groups. In the younger age group IL-10/pro-inflammatory cytokine ratios were significantly higher and IL-10-skewed (P= 0.0207 and P= 0.0111 for IL-10:TNF-α and IL-10:IFN-γ ratios, respectively) in the those developing uncomplicated malaria compared to those with persistent asymptomatic malaria in younger age group (**Figure 5B**). In this group those with persistent asymptomatic malaria had a ratios of 1.001±0.1355 for IL-10:TNF-α and 1.016±0.1794 for IL-10:IFN-γ) displaying an even balance of IL-10: proinflammatory cytokine. On the other hand adults did not have IL-10 skewed ratios with TNF-α or IFN-γ suggesting that a different or additional mechanism may facilitate the persistence of asymptomatic malaria in the older age group. Collectively this data suggests that IL-10 is not sufficient to dampen inflammatory responses in *Plasmodium* infection that lead to the fevers that define uncomplicated malaria.

## Discussion

Significant gaps exist in the understanding of the contribution of anti-parasite and anti-disease immunity in the persistent asymptomatic parasite carriage in areas with high transmission of *P. falciparum*. Mathematical modelling of existing data indicates that asymptomatic *P. falciparum* infections could last for as long as 13-years [40] and field data documents the longevity of asymptomatic infections ranges from at least 2-9 months [7,41,42]. As we have previously published, our study area has high rate of persistent asymptomatic parasite carriage over the 3 month time period of study [24]. However, a proportion of individuals in our study converted to symptomatic malaria as defined by the development of fever. Thus, we have assessed immune correlates of asymptomatic carriage of *P. falciparum*, as well as the predictors of conversion to symptomatic malaria.

Consistent with the acquisition of immune response with age and exposure [13,43] in our study, older asymptomatic individuals had better control of circulating iRBCs with a greater proportion of sub-microscopic infections than younger individuals. This was associated with an increase in the quantity of circulating anti-parasite IgG which appeared to saturate at around 20 years old. Our data is in agreement with an age-structured mathematical model of malaria transmission which showed that anti-parasite immunity which results in more rapid clearance of parasitemia has an approximate half-life of 20 years [44]. With a focus on the younger age group, there did not appear to be any correlations with respect to parasitemia levels and IgG levels or the avidity of the IgG. The *in vitro* invasion-blocking capability of the plasma in young individuals had some functionality with respect to invasion blocking, but only at higher parasitemia levels. In this assay plasma factors other than IgG, such as IgM [45,46], may contribute to this observation and control of *P. falciparum* parasitemia in children. Collectively these data support a role for IgG in exerting control of iRBCs in *P. falciparum* infections, but it may not be the principal factor that dictates asymptomatic carriage of parasites in younger individuals.

Surprisingly the proportion of those developing uncomplicated malaria over the course of our study was comparable between the younger and the older age groups, suggesting that the conversion from an asymptomatic to symptomatic infection is independent of age in highly exposed individuals. The reasons for this finding are not clear but may be related to heterogeneity of the circulating *P. falciparum* parasite population and emergence of highly inflammatory parasite clonotypes [47,48] that overcome the pyrogenic threshold of individuals. Consistent with the oscillation of parasite densities over time reported in a Vietnam cohort [7], parasitemia in persistent asymptomatic infections in our study fluctuated widely and independently of IgG levels, and different clonotypes of *P. falciparum* were observed at different time points in this study.

Cytokine profiles are considerably more dynamic than IgG responses. In line with the hypothesis that clinical immunity to malaria is mediated by a tempering of inflammation, previous studies have shown that the clinical outcome of malaria infection is largely dictated by the balance between pro-inflammatory and anti-inflammatory markers [17,18]. Although our study highlighted that carriage of *P. falciparum* across the duration of this study was associated with a consistent inflammatory response characterized by TNF-α, IL-10 and MCP-1, we found no obvious cytokine signature associated with the asymptomatic carriage of *P. falciparum* when compared to symptomatic individuals. Similar to findings in a study of visceral leishmaniasis [49], where increased IL-10 levels were the primary predictor of clinical conversion from asymptomatic to symptomatic visceral leishmaniasis, the conversion from asymptomatic to symptomatic malaria in our study was linked to increased baseline levels of IL-10 skewed ratios with TNF-α and IFN-γ in children at the initial time point of our study.

Although IL-10 is known to exert anti-inflammatory effects that include inhibition of monocytes and macrophages in the production of inflammatory cytokines such as IL-6, TNF-α, and IL-l [50], in agreement with a recent study from the Gambia [51] in our study IL-10 was associated with the presence of *Plasmodium* parasites. Given that higher levels of IL-6, IL-1RA and MCP-1 were also associated with symptomatic infections in the younger age group of our study, IL-10 levels are likely a consequence of increased inflammation. Notably, IL-10 induces IL1RA, a potent anti-inflammatory cytokine that regulates the pro-inflammatory actions of IL-1 [52] that has previously been shown to be associated with severity of malarial disease [53]. At the baseline time point in our study, we found that balanced inflammatory cytokine ratios were correlated with developing symptoms from *P. falciparum* infection in our younger age group. Specifically, those who developed uncomplicated malaria had a significant IL-10 skewed ratio at the initial time point compared to those who had persistent asymptomatic malaria. An IL-10 skewed ratio may indicate that an individual will shortly develop symptoms of *P. falciparum* infection.

Most of the association between biomarkers and clinical malaria in this study were only found in the younger age group, but not in adults, indicating that the mechanisms of the protection against clinical malaria may be different according to age. As such, more studies into how asymptomatic carriage of *Plasmodium* occurs are warranted and should include non-antibody mediated mechanisms of parasite control and clinical immunity which may be relevant in children.

## Data Availability

All data produced in the present study are available upon reasonable request to the authors

## Acknowledgements

We thank the field staff and, most importantly, residents of the Esse Health district in Cameroon who participated in this study.

## Author contributions

BF devised the experiments, performed the experiments, analyzed data, and wrote the paper; FM, MFA, EE, CD, GC, MK, RK and SK performed fieldwork and sample processing; MS and RP provided cytokine kits and recombinant *P. falciparum* antigens, respectively and reviewed paper. RM supervised the experiments and reviewed paper; LA and TL devised the experiments, supervised the experiments, analyzed data, and wrote the paper.

