## Supplementary file for "Asymptomatic carriage of *Plasmodium falciparum* in children living in a hyperendemic area occurs independently of IgG responses but is associated with a balanced inflammatory cytokine ratio"


### AUTHORS AND AFFILIATIONS

Balotin Fogang[1,2], Matthieu Schoenhals[1], Franklin M. Maloba[1], Marie Florence Biabi[1,3], Estelle Essangui[1], Christiane Donkeu[1,2], Glwadys Cheteug[1,4], Marie Kapen[1], Rodrigue Keumoe[1], Sylvie Kemleu[1], Sandrine Nsango[1,5], Douglas H. Cornwall[6], Carole Eboumbou[1,5], Ronald Perraut[1], Rosette Megnekou[2], Tracey J. Lamb*[6], Lawrence S. Ayong*[1]

**Affiliations:**

[1] Molecular Parasitology Laboratory, Centre Pasteur du Cameroun, BP 1274 Yaounde, Cameroon,

[2] Department of Animal Biology and Physiology of the University of Yaoundé I, BP 812 Yaounde, Cameroon,

[3] Department of Biochemistry, University of Douala, BP 24157 Douala, Cameroon,

[4] Department of Medical Laboratory Sciences, University of Buea, BP 63 Buea, Cameroon

[5]Faculty of Medicine and Pharmaceutical Sciences, University of Douala, BP 2701 Douala, Cameroon

[6] Department of Pathology, University of Utah, 15 N Medical Drive, Salt Lake City 84112, USA.

* Co-corresponding authors

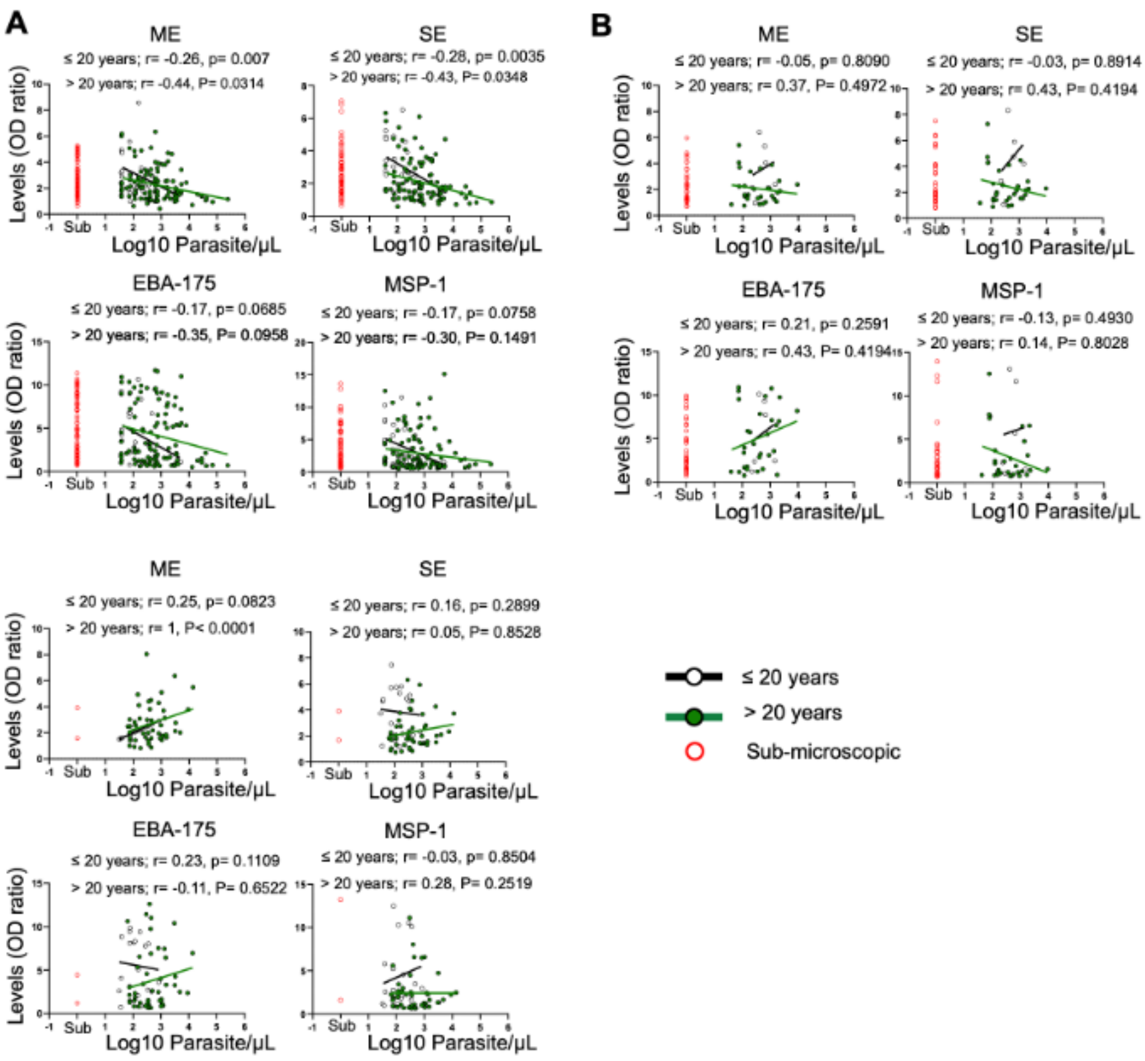


**Supplementary Figure 1. Correlation between IgG anti-*Plasmodium* antibody levels and parasite loads.** Scatterplots show the correlation between baseline IgG anti-*P. falciparum* antibody levels and parasitemia at the detection limit of microscopy in different age groups. Spearman rank correlation is presented as the best fit line and the coefficient (r), as well as the p-values (p), are shown for each age group. SE: Schizont extract. ME: Merozoite extract. EBA-175: Erythrocyte binding antigen-175. MSP-1: Merozoite surface protein-1. OD: optical density.

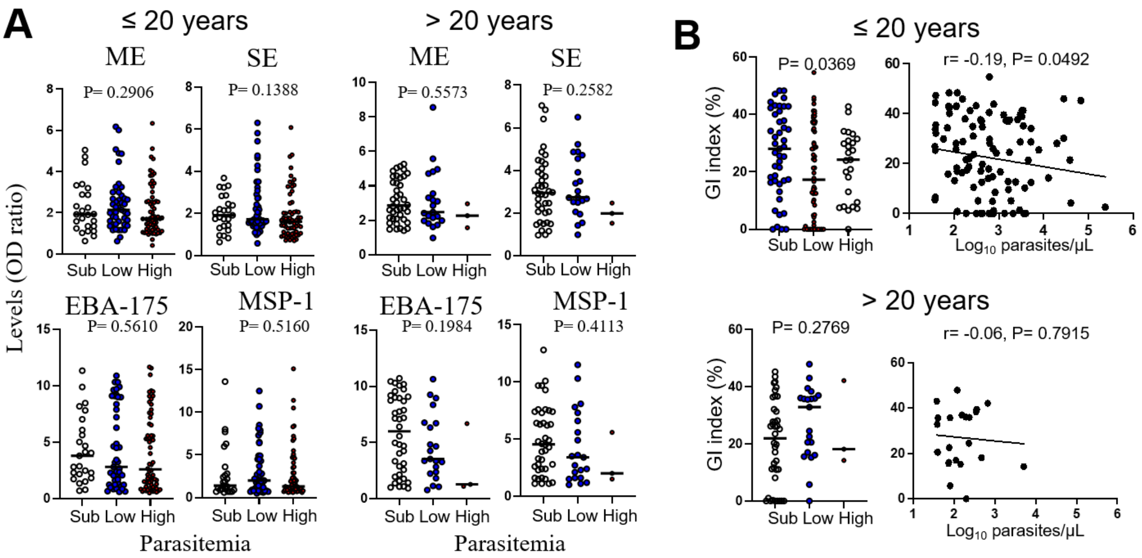


**Supplementary Figure 2. IgG anti-*Plasmodium* antibody levels (A) and anti-parasite growth inhibition activities (B) at baseline divided according to parasitemia levels.** A) Data presented in the scatterplots represent IgG antibody levels against different *P. falciparum* antigens (ME, SE, EBA-175 and MSP-1) in parasitemic groups, according to age groups collected at the baseline of the study. Median levels of parasitemia between the three groups were compared by a Kruskal-Wallis test and the p-values are shown on the plot. None were significant and therefore no two-way comparisons were performed. B) The scatterplots on the left show *P. falciparum* growth inhibition activities of circulating plasma antibodies in parasitemic groups. Median antibody GI index between parasitemia the three groups were compared by Kruskal-Wallis test and the p-values are shown on the plot. The scatterplots on the right show the correlation between *P. falciparum* growth inhibition activities of circulating antibodies and parasitemia in different age groups. Spearman rank correlation is presented as the best fit line and the coefficient (r), as well as the p-values (p), are shown for each age group. Parasite densities were groups in three groups as follows, submicroscopic parasitemia, had low levels of microscopic parasitemia (parasitemia ≤ 413.1 parasites/µl, representing the median parasitemia in the study population) and high levels of parasitemia (parasitemia> 413.1 parasites/µl). SE: Schizont extract. ME: Merozoite extract. EBA-175: Erythrocyte binding antigen-175. MSP-1: Merozoite surface protein-1. GI: Growth inhibition. OD: optical density. Sub: Submicroscopic. L

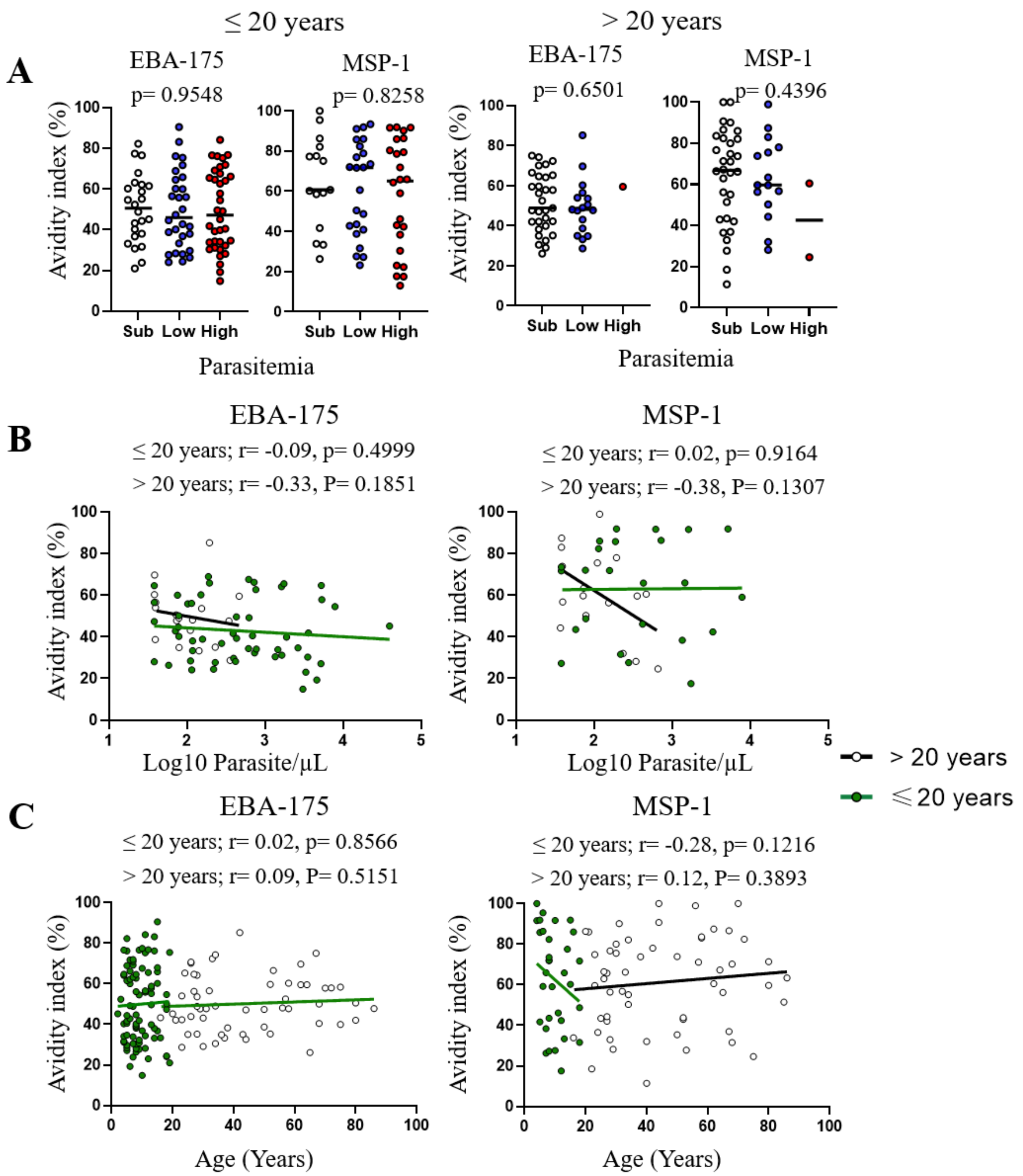


**Supplementary Figure 3. No association was found between IgG anti-*Plasmodium* antibody avidity and parasite densities.** A) Data presented in the scatterplots represent IgG antibody avidity index against different *P. falciparum* antigens (EBA-175 and MSP-1) in individuals with microscopic parasitemia at the baseline, according to age groups. Median levels between parasitemia the three groups were compared by Kruskal-Wallis test and the p-values are shown on the plot. In scatterplots show the correlation between IgG anti-*Plasmodium* antibody avidity index and parasitemia. Parasite densities were groups in three groups as follows, submicroscopic parasitemia, had low levels of microscopic parasitemia (parasitemia ≤ 413.1 parasites/μl, representing the median parasitemia in the study population) and high levels of parasitemia (parasitemia> 413.1 parasites/μl). Spearman rank correlation is presented as the best fit line and the coefficient (r) and the p-values (p), are shown for each age group. EBA-175: Erythrocyte binding antigen-175. MSP-1: Merozoite surface protein-1.

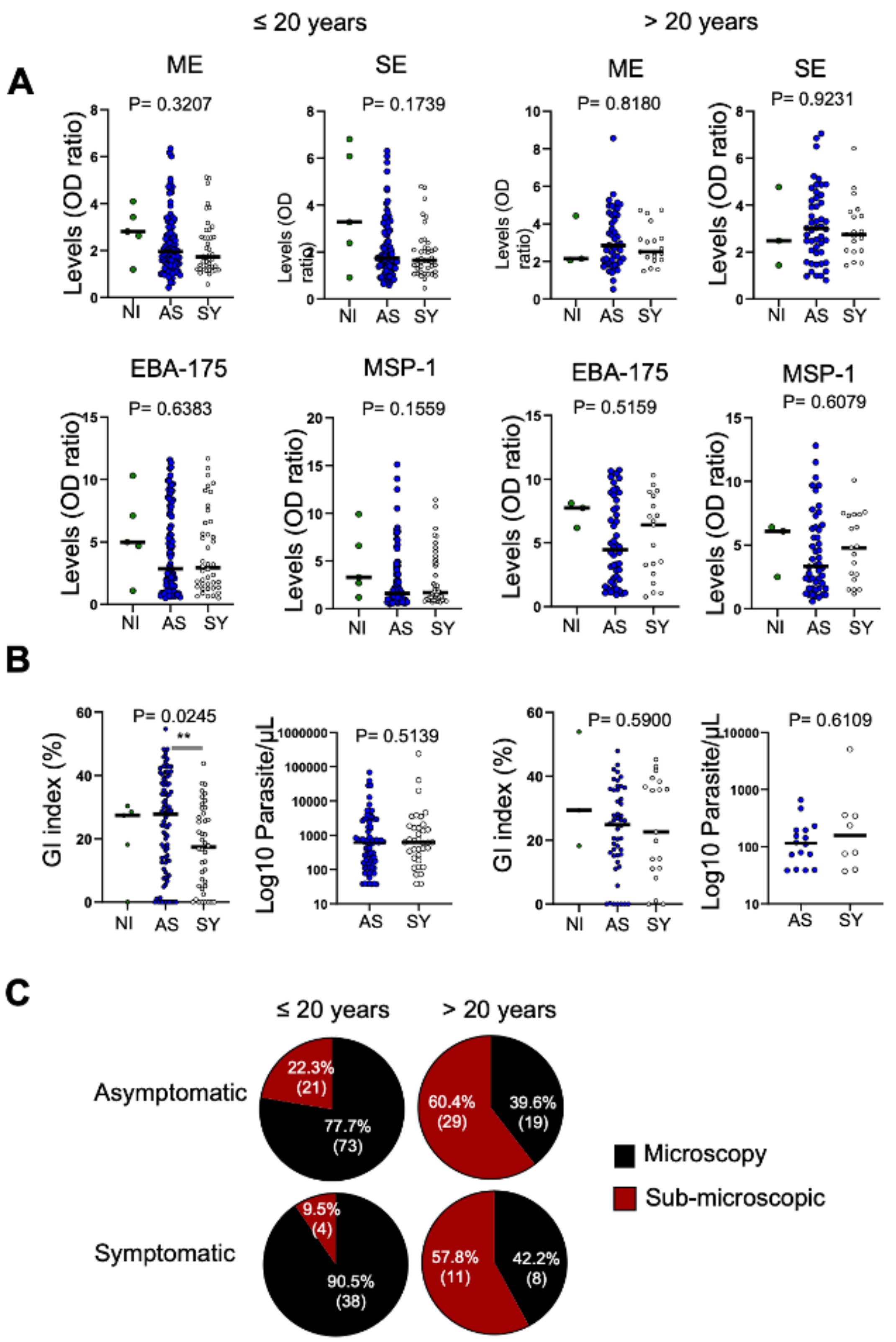


**Supplementary Figure 4. No association was found between IgG anti-*Plasmodium* antibody levels and infection status.** A) Data presented in the scatterplots represent baseline

IgG antibody levels against different *P. falciparum* antigens (ME, SE, EBA-175 and MSP-1) by infection status, according to age group. B) scatterplots represent the parasite growth inhibition index of circulating antibody or microscopic parasite densities in the study population groups at the baseline. Median levels were compared using nonparametric Kruskal-Wallis test for multiple comparisons and Wilcoxon signed-rank test for comparison between two groups. In Panel A, pairwise comparisons were performed only when multiple comparisons were significant at $p<0.05$. SE: Schizont extract. ME: Merozoite extract. EBA-175: Erythrocyte binding antigen-175. MSP-1: Merozoite surface protein-1. NI: Non-infected. AS: Asymptomatic. SY: Symptomatic. GI: Growth inhibition. OD: optical density. **p< 0.05.

**Supplementary Table 1. Multivariate logistic regression analysis of selected cytokines**

| Cytokines | Coefficients | P value |
|---|---|---|
| IL-10 | -1.19 | 0.1828 |
| **IL-1RA** | **2.18** | **0.0208** |
| MCP-1 | 1.80 | 0.1026 |
| IP-10 | 0.87 | 0.2558 |
| IL-6 | -0.35 | 0.5888 |
| **IL-10/TNF-α** | **2.28** | **0.0130** |

The data in this table represents the results of multivariate logistic regression analysis of selected cytokines with p-value <0.1 when comparing individuals with asymptomatic *Plasmodium* infection to those with symptomatic infection.
